# Real-world effectiveness of nirsevimab over three RSV seasons in an integrated health care system: a target trial emulation study

**DOI:** 10.64898/2026.11.12.26367940

**Authors:** Sara S Kim, Emily Wu, Alexander F Hudgins, Mackenzie Crawford, Marina Bruck, Felipe Lobelo, David Benkeser, Elizabeth T Rogawski McQuade

## Abstract

Most nirsevimab effectiveness estimates come from test-negative designs, which restrict populations to those seeking healthcare and testing. We estimated nirsevimab effectiveness among infants six months post-receipt using a target trial emulation with an estimand aligned to phase 3 trials.

We emulated a target trial among infants born from February 2, 2023 through May 30, 2026, using electronic medical records at Kaiser Permanente Georgia. Using a g-computation estimator with Cox models fit separately in nirsevimab recipients and non-recipients, we estimated the cumulative incidence of RSV hospitalization and medically-attended RSV at 30-day intervals through 180 days post-receipt.

Among 4,826 infants, nirsevimab reduced the risk of RSV hospitalization by 95% (95% CI: 84% to 98%) and medically-attended RSV by 67% (95% CI: 45% to 80%) at 180 days post-receipt. The number needed to immunize to prevent one case was 147 (95% CI: 94 to 333) for RSV hospitalization and 39 (95% CI: 28 to 68) for medically-attended RSV.

Nirsevimab was effective through 180 days post-receipt. This cohort-based emulation avoids conditioning on testing and the restricted generalizability of the test-negative and matching-based designs that dominate the literature. These findings support continued nirsevimab use to protect infants against RSV.

## Introduction

Respiratory syncytial virus (RSV) is the leading cause of infant hospitalization in the United States (U.S.).^1^ Among U.S. children aged less than five years, an estimated 2.1 million children have an RSV-related medically-attended illness annually.^2,3^ Most children (97%) are expected to be infected by their second birthday.^4^ RSV generally follows a winter seasonality with activity between October and March.^5,6^ Because treatment of RSV illness in infants remains largely supportive, prevention is the main lever for reducing burden.^7–9^

Since 2023, the Advisory Committee on Immunization Practices (ACIP) has recommended that all infants be protected through either maternal RSV vaccination (RSVpreF) during pregnancy or a long-acting monoclonal antibody given to the infant. Nirsevimab and, since June 2025, clesrovimab are recommended without preference for infants aged less than eight months who are born during or entering their first RSV season and whose mothers were not vaccinated.^10–12^ Across three randomized clinical trials (RCTs),^13–15^ nirsevimab reduced the risk of hospitalization for RSV-associated lower respiratory tract infection (LRTI) by 77% (95% confidence interval [CI]: 50% to 90%) and medically-attended RSV-associated LRTI by 80% (95% CI: 66% to 88%) at 150 days post intervention.^16^ In trials, clesrovimab reduced the risk of RSV-associated hospitalization by 84% (95% CI: 66% to 93%) and medically-attended RSV LRTI by 60% (95% CI: 44% to 72%) through day 150 post intervention.^17^ Several U.S.-based post-licensure effectiveness studies have demonstrated that nirsevimab was highly effective against RSV-related medically-attended illness,^18–24^ hospitalization,^18,19,21–26^ and intensive care unit admission^27^ during the 2023-2024 and 2024-2025 seasons. Pooled estimates from more than 40 observational studies, most of them European and nearly all test-negative or matched-cohort designs, put effectiveness against RSV hospitalization at roughly 80–85%, with similar protection against intensive care admission.^7,28–30^

Most observational studies evaluating RSV preventive interventions have used the test-negative design (TND).^18–22,24,25,27^ While TND is efficient and can control for healthcare seeking and testing behavior,^31,32^ bias may arise when effectiveness estimates condition on individuals who have been tested and when the intervention impacts healthcare seeking and testing behavior.^33^ Additionally, estimands in TND studies are less clearly defined, particularly with respect to the target population of interest and the timing of intervention receipt. For example, when nirsevimab is administered at different times relative to the RSV season or an infant’s age, an estimate may include infants with very different follow-up windows and baseline risk. Surveillance-based TND estimates from Spain for 2024–2025 showed strong age-period-cohort effects and an early apparent decline in effectiveness that the authors attributed in part to these design features.^34^ Finally, because TND studies do not estimate risks, their estimates cannot be directly compared to those reported in RCTs or used to estimate the absolute burden of disease prevented.

In addition to the ability to estimate absolute risks, applying a target trial emulation (TTE) framework within a cohort study design may overcome some of the limitations of TND studies by including a more generalizable population and more explicitly defined estimand.^35–37^ This framework can mitigate confounding and selection bias, avoid conditioning on testing, and establish a consistent time zero at which follow-up begins for both nirsevimab recipients and non-recipients.

Matching is one of the most common TTE approaches.^38^ In traditional matching, intervention recipients are matched to individuals with no intervention based on a selected set of confounding covariates.^35^ Follow-up for the pair is then initiated at the time of nirsevimab administration for the recipient, defining a comparable time zero for the individual receiving no intervention. The recent nationwide French evaluations of nirsevimab and of maternal RSVpreF used this matched approach.^39,40^ However, individuals without a matching counterpart are excluded from the study, potentially limiting efficiency and generalizability.^35,41^ Therefore, we applied a novel TTE method that targets a similar estimand to matching-based studies but uses data from all observed individuals without requiring explicit matching to define time zero.^42^

To address the potential limitations of estimates derived from prior nirsevimab studies, we constructed a birth cohort using electronic medical records (EMRs) from an integrated healthcare system. We evaluated the effectiveness of nirsevimab against RSV hospitalization and medically-attended RSV compared to no intervention 30 to 180 days after nirsevimab receipt. Our effectiveness estimates covered the first three RSV seasons during which nirsevimab was available in the U.S.

## Methods

### Setting and data source

This study included members from Kaiser Permanente Georgia (KPGA), an integrated healthcare delivery system serving more than 400,000 insured members in metropolitan Atlanta and North Georgia. KPGA delivers primary, pediatric, specialty, and urgent care in its own medical offices, while emergency department and inpatient care are provided at contracted community hospitals and captured through claims. KPGA has an extensive data repository of EMRs, including information related to patient demographics, diagnoses, procedures, claims, laboratory values, and prescribed medications. The data for this study were pulled on June 16, 2026. The Kaiser Permanente Institutional Review Board approved the study (IRB #2261961) with a waiver of informed consent given minimal risk to participants.

### Study design

We aimed to emulate a hypothetical trial in which infants are randomized to either nirsevimab or placebo to estimate the effectiveness of first nirsevimab dose compared to no intervention during the 180 days from nirsevimab receipt (Table S1). We constructed an infant-mother paired birth cohort and followed infants from birth until the first of an outcome of interest or right censoring event. Right-censoring events were a lapse in coverage of more than 75 days, the end of the follow-up window (180 days from nirsevimab receipt), the study end date (May 30, 2026), or nirsevimab receipt among the unexposed.

### Eligibility

We included infants born into KPGA between February 2, 2023 (aged <8 months on October 1, 2023) and May 30, 2026. We excluded infants if they could not be paired with their mother’s record, received palivizumab or an unknown RSV product, their mother received the bivalent prefusion F RSV (RSVpreF) vaccination ≥14 days before delivery, or were not enrolled into KPGA within 75 days from birth. Newborns were automatically covered under the mother’s plan for the first 31 days of life. Allowing one additional day as an administrative buffer, we considered infants enrolled by day 32 to have continuous coverage from birth. For infants enrolled between 33 and 75 days from birth, we additionally required a documented healthcare encounter at KPGA within 75 days from birth to ensure they were not enrolled elsewhere.

### Exposure

The exposure of interest was receipt of first nirsevimab dose. We did not restrict at what age infants received the first dose. Additionally, we included nirsevimab recipients whose mothers received maternal RSV vaccination <14 days prior to delivery. We assumed protection began immediately after nirsevimab receipt. We also evaluated a seven-day lag in protection from receipt.

We identified nirsevimab receipt in the infant’s medical record using the Centers for Disease Control and Prevention’s (CDC) standardized vaccine code sets. We identified RSV preventive products using immunization product codes (CVX) and product administration codes (CPT) linked to the CDC’s predefined vaccine code set for RSV. We defined nirsevimab receipt by CPT codes 90380 and 90381 and CVX codes 306 and 307; administrations recorded only with non-specific RSV product codes (CVX 304, 314, or 315) were classified as unknown RSV product.

To determine maternal RSV vaccination timing in relation to infant delivery, we used CPT code 90678 and CVX code 305. We linked maternal vaccinations to their respective infants using a distinct pregnancy identification (ID). We determined this linkage using the pregnancy outcome date and infant date of birth. For multiple births, one pregnancy ID could link to more than one infant.

### Outcome

The primary outcomes were the first RSV hospitalization and the first medically-attended RSV event occurring between birth and the end of follow up. Medically-attended RSV included virtual care, outpatient, emergency department, and inpatient visits. We identified RSV-related encounters using International Classification of Diseases, 10^th^ Revision (ICD-10) codes for RSV from the infant’s EMR. We pulled diagnoses and corresponding encounters for infants where the ICD-10 code was flagged as an acute respiratory infection (Appendix A). We classified encounters with RSV-specific ICD-10 codes (J21.0, B97.4, J12.1, J20.5) as medically-attended RSV encounters. We considered codes in any diagnosis position. We identified hospitalizations from inpatient claims from contracted hospitals with a relevant diagnosis code.

Secondary outcomes included the first medically-attended all-cause bronchiolitis event, all-cause bronchiolitis hospitalization, medically-attended all-cause acute respiratory illness (ARI), and all-cause ARI hospitalization. We identified bronchiolitis encounters with bronchiolitis specific ICD-10 codes (J21.0, J21.1, J21.8, J21.9) and all-cause ARI encounters with ICD-10 codes from Appendix A.

### Statistical analysis

To estimate nirsevimab effectiveness, we estimated cumulative incidence of the outcome of interest with and without nirsevimab at 30-day intervals through 180 days from nirsevimab receipt by using a g-computation-based estimator. This estimator included separate Cox proportional hazards models with age as the time scale to model risk of outcomes among infants after receipt of nirsevimab, as well as risk among children who had not yet received nirsevimab.^42^ We marginalized over infant age and covariates including maternal age at delivery, number of prenatal encounters during the relevant pregnancy, maternal Tdap and influenza vaccination receipt, race, calendar time using harmonic terms to account for seasonality, and infant complications including preterm birth, congenital malformations of the circulatory system, congenital pulmonary abnormalities, and neuromuscular conditions.^43–47^

We defined effectiveness or relative risk reduction (RRR) as (1 − risk ratio [RR]) × 100% and estimated the risk difference (RD). Additionally, we estimated the number needed to immunize (NNI) as the absolute value of 1/RD to indicate the number of infants who would need to receive nirsevimab to prevent one additional case. We generated 95% confidence intervals using 1000 nonparametric bootstrap samples. When the associated RD 95% CIs crossed zero, we reported discontinuous intervals for NNI.^48^

We conducted all analyses in R v4.4.2. To obtain the cumulative incidence estimates and accompanying RRR and RD using the novel statistical approach, we used the ‘*nomatch*’ package v0.1.0.^49^

## Results

Between February 2, 2023, and May 30, 2026, 10,727 infants were born into KPGA. We excluded infants who were not enrolled into KPGA within 75 days from birth (n=4,126), who could not be linked to their mother’s record (n=894), whose mothers received maternal RSV vaccination ³14 days before delivery (n=873), and who received palivizumab (n=6) or unknown RSV preventive product (n=2) for a total of 4,826 mother-infant pairs included in analysis (Figure S1).

Over three RSV seasons, 1,858 (38%) infants received nirsevimab. The proportion of infants who received nirsevimab increased over time. Among infants born prior to July 2023, 4% received nirsevimab, reflecting the national supply shortage and CDC’s interim prioritization guidance that season.^43^ Among infants born in the 2023-2024, 2024-2025, and 2025-2026 birth cohorts, 40%, 50%, and 48% received nirsevimab, respectively. Infants who did and did not receive nirsevimab had similar distributions of characteristics, including maternal age, number of prenatal care encounters during pregnancy, maternal comorbidities during pregnancy, and newborn complications (Table 1). However, infants who had mothers receiving other maternal vaccinations such as for Tdap or influenza were more likely to receive nirsevimab. Moreover, Black or African American infants were less likely to receive nirsevimab compared to infants of other races (36% versus 45% among Asian infants and 43% among White infants).

**Table 1.**
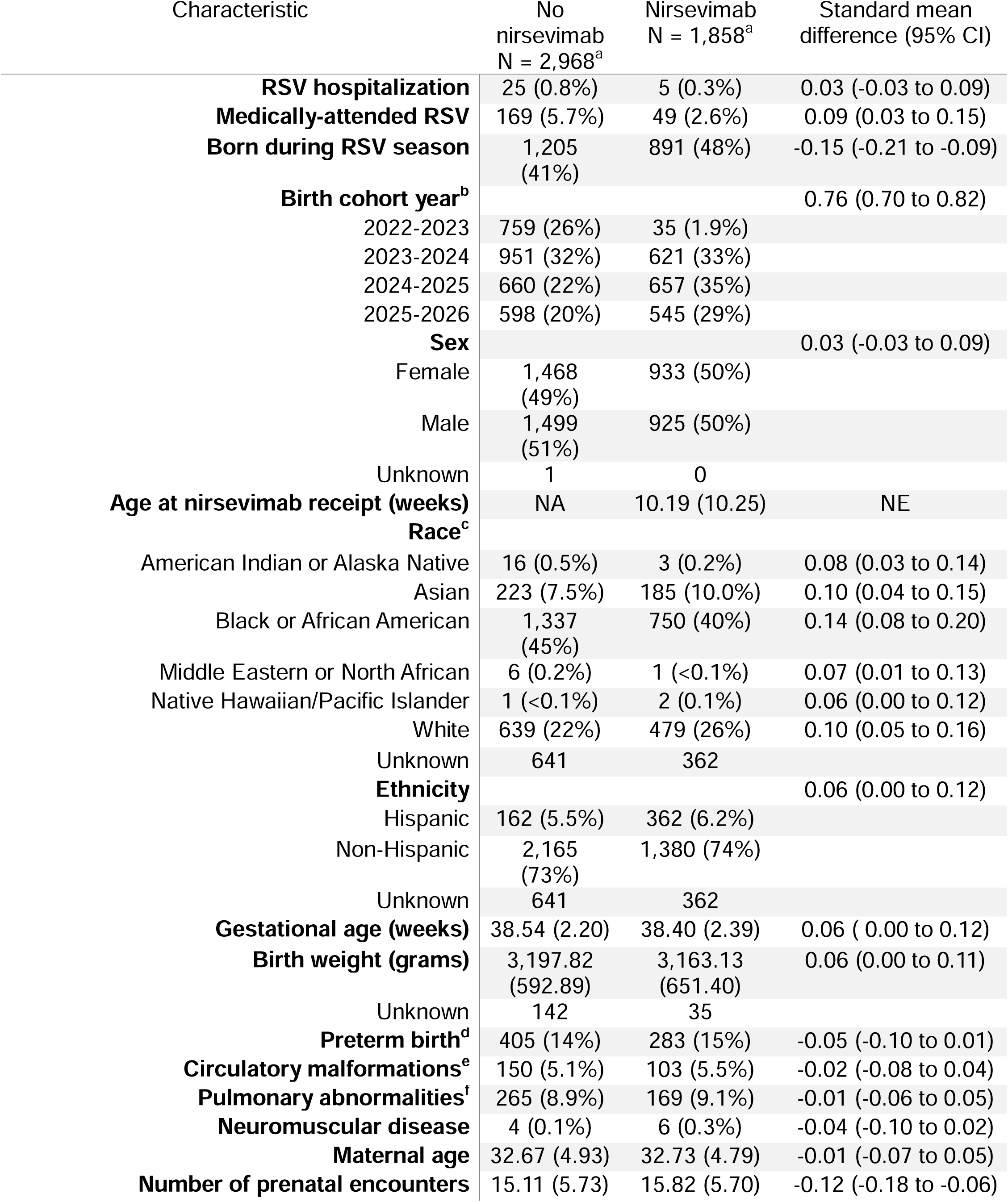
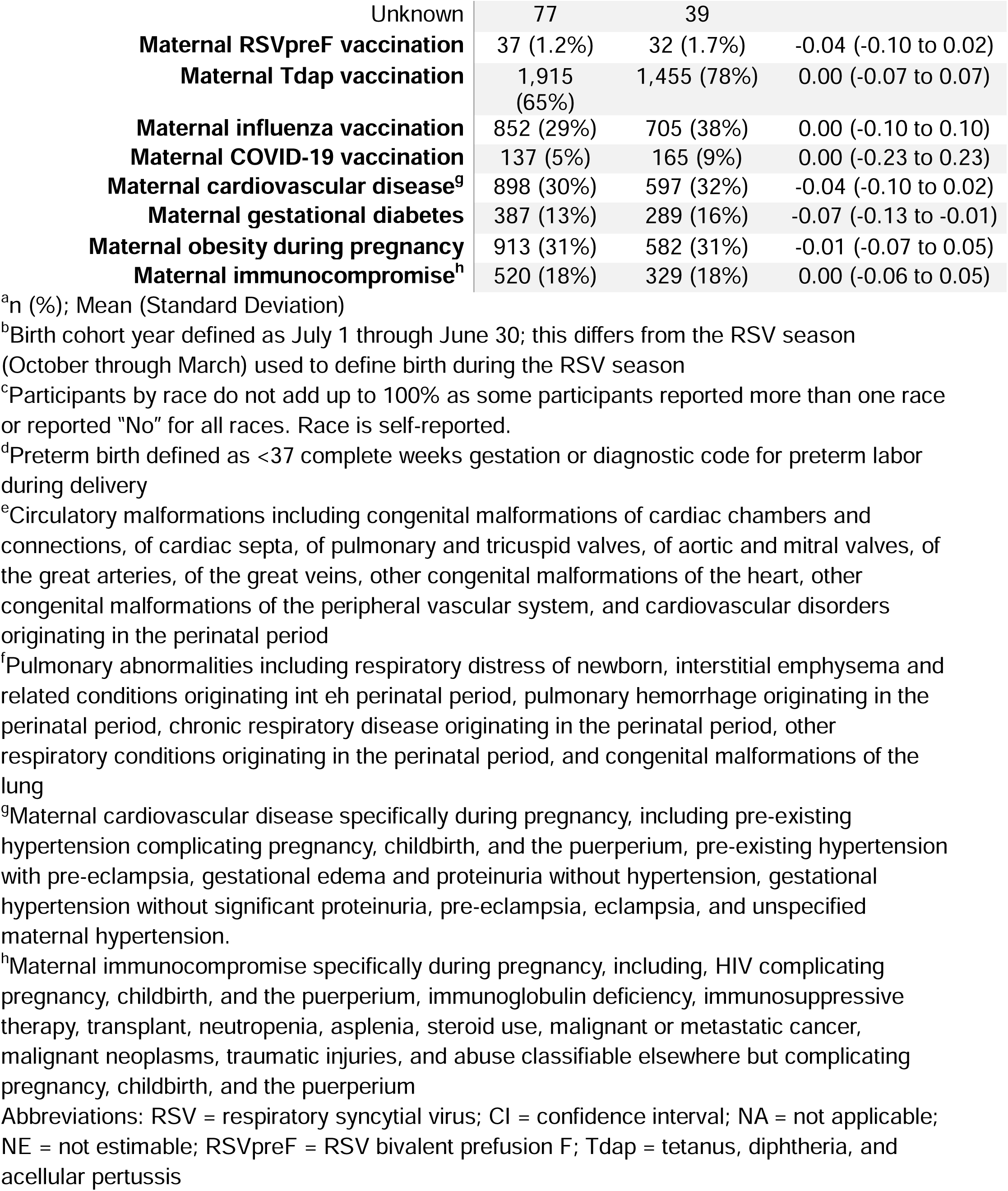
Infant and maternal characteristics by nirsevimab receipt.

| Characteristic | No nirsevimab<br>N = 2,968<sup>a</sup> | Nirsevimab<br>N = 1,858<sup>a</sup> | Standard mean difference (95% CI) |
|---|---|---|---|
| **RSV hospitalization** | 25 (0.8%) | 5 (0.3%) | 0.03 (-0.03 to 0.09) |
| **Medically-attended RSV** | 169 (5.7%) | 49 (2.6%) | 0.09 (0.03 to 0.15) |
| **Born during RSV season** | 1,205 (41%) | 891 (48%) | -0.15 (-0.21 to -0.09) |
| **Birth cohort year<sup>b</sup>** |  |  | 0.76 (0.70 to 0.82) |
| 2022-2023 | 759 (26%) | 35 (1.9%) |  |
| 2023-2024 | 951 (32%) | 621 (33%) |  |
| 2024-2025 | 660 (22%) | 657 (35%) |  |
| 2025-2026 | 598 (20%) | 545 (29%) |  |
| **Sex** |  |  | 0.03 (-0.03 to 0.09) |
| Female | 1,468 (49%) | 933 (50%) |  |
| Male | 1,499 (51%) | 925 (50%) |  |
| Unknown | 1 | 0 |  |
| **Age at nirsevimab receipt (weeks)** | NA | 10.19 (10.25) | NE |
| **Race<sup>c</sup>** |  |  |  |
| American Indian or Alaska Native | 16 (0.5%) | 3 (0.2%) | 0.08 (0.03 to 0.14) |
| Asian | 223 (7.5%) | 185 (10.0%) | 0.10 (0.04 to 0.15) |
| Black or African American | 1,337 (45%) | 750 (40%) | 0.14 (0.08 to 0.20) |
| Middle Eastern or North African | 6 (0.2%) | 1 (<0.1%) | 0.07 (0.01 to 0.13) |
| Native Hawaiian/Pacific Islander | 1 (<0.1%) | 2 (0.1%) | 0.06 (0.00 to 0.12) |
| White | 639 (22%) | 479 (26%) | 0.10 (0.05 to 0.16) |
| Unknown | 641 | 362 |  |
| **Ethnicity** |  |  | 0.06 (0.00 to 0.12) |
| Hispanic | 162 (5.5%) | 362 (6.2%) |  |
| Non-Hispanic | 2,165 (73%) | 1,380 (74%) |  |
| Unknown | 641 | 362 |  |
| **Gestational age (weeks)** | 38.54 (2.20) | 38.40 (2.39) | 0.06 ( 0.00 to 0.12) |
| **Birth weight (grams)** | 3,197.82 (592.89) | 3,163.13 (651.40) | 0.06 (0.00 to 0.11) |
| Unknown | 142 | 35 |  |
| **Preterm birth<sup>d</sup>** | 405 (14%) | 283 (15%) | -0.05 (-0.10 to 0.01) |
| **Circulatory malformations<sup>e</sup>** | 150 (5.1%) | 103 (5.5%) | -0.02 (-0.08 to 0.04) |
| **Pulmonary abnormalities<sup>f</sup>** | 265 (8.9%) | 169 (9.1%) | -0.01 (-0.06 to 0.05) |
| **Neuromuscular disease** | 4 (0.1%) | 6 (0.3%) | -0.04 (-0.10 to 0.02) |
| **Maternal age** | 32.67 (4.93) | 32.73 (4.79) | -0.01 (-0.07 to 0.05) |
| **Number of prenatal encounters** | 15.11 (5.73) | 15.82 (5.70) | -0.12 (-0.18 to -0.06) |
| Unknown | 77 | 39 |  |
| **Maternal RSVpreF vaccination** | 37 (1.2%) | 32 (1.7%) | -0.04 (-0.10 to 0.02) |
| **Maternal Tdap vaccination** | 1,915 (65%) | 1,455 (78%) | 0.00 (-0.07 to 0.07) |
| **Maternal influenza vaccination** | 852 (29%) | 705 (38%) | 0.00 (-0.10 to 0.10) |
| **Maternal COVID-19 vaccination** | 137 (5%) | 165 (9%) | 0.00 (-0.23 to 0.23) |
| **Maternal cardiovascular disease<sup>g</sup>** | 898 (30%) | 597 (32%) | -0.04 (-0.10 to 0.02) |
| **Maternal gestational diabetes** | 387 (13%) | 289 (16%) | -0.07 (-0.13 to -0.01) |
| **Maternal obesity during pregnancy** | 913 (31%) | 582 (31%) | -0.01 (-0.07 to 0.05) |
| **Maternal immunocompromise<sup>h</sup>** | 520 (18%) | 329 (18%) | 0.00 (-0.06 to 0.05) |

<sup>a</sup>n (%); Mean (Standard Deviation)
<sup>b</sup>Birth cohort year defined as July 1 through June 30; this differs from the RSV season (October through March) used to define birth during the RSV season
<sup>c</sup>Participants by race do not add up to 100% as some participants reported more than one race or reported “No” for all races. Race is self-reported.
<sup>d</sup>Preterm birth defined as <37 complete weeks gestation or diagnostic code for preterm labor during delivery
<sup>e</sup>Circulatory malformations including congenital malformations of cardiac chambers and connections, of cardiac septa, of pulmonary and tricuspid valves, of aortic and mitral valves, of the great arteries, of the great veins, other congenital malformations of the heart, other congenital malformations of the peripheral vascular system, and cardiovascular disorders originating in the perinatal period
<sup>f</sup>Pulmonary abnormalities including respiratory distress of newborn, interstitial emphysema and related conditions originating int eh perinatal period, pulmonary hemorrhage originating in the perinatal period, chronic respiratory disease originating in the perinatal period, other respiratory conditions originating in the perinatal period, and congenital malformations of the lung
<sup>g</sup>Maternal cardiovascular disease specifically during pregnancy, including pre-existing hypertension complicating pregnancy, childbirth, and the puerperium, pre-existing hypertension with pre-eclampsia, gestational edema and proteinuria without hypertension, gestational hypertension without significant proteinuria, pre-eclampsia, eclampsia, and unspecified maternal hypertension.
<sup>h</sup>Maternal immunocompromise specifically during pregnancy, including, HIV complicating pregnancy, childbirth, and the puerperium, immunoglobulin deficiency, immunosuppressive therapy, transplant, neutropenia, asplenia, steroid use, malignant or metastatic cancer, malignant neoplasms, traumatic injuries, and abuse classifiable elsewhere but complicating pregnancy, childbirth, and the puerperium
Abbreviations: RSV = respiratory syncytial virus; CI = confidence interval; NA = not applicable; NE = not estimable; RSVpreF = RSV bivalent prefusion F; Tdap = tetanus, diphtheria, and acellular pertussis

Infants born out of season received nirsevimab later in life because they were not eligible until the start of their first season (Figure 1). Among infants born during the RSV season, the median number of days between birth and nirsevimab receipt was 7 days (Interquartile Range (IQR): 4 to 22). Among infants born outside of the RSV season, the median number of days between birth and nirsevimab receipt was 124 days (IQR: 63 to 182). All but 9 infants (<1%) received their first nirsevimab dose within eight months from birth, aligning with the general recommendations for nirsevimab receipt. Among 69 infants whose mothers received RSV vaccination <14 days from delivery, 46% received nirsevimab.

**Figure 1.**
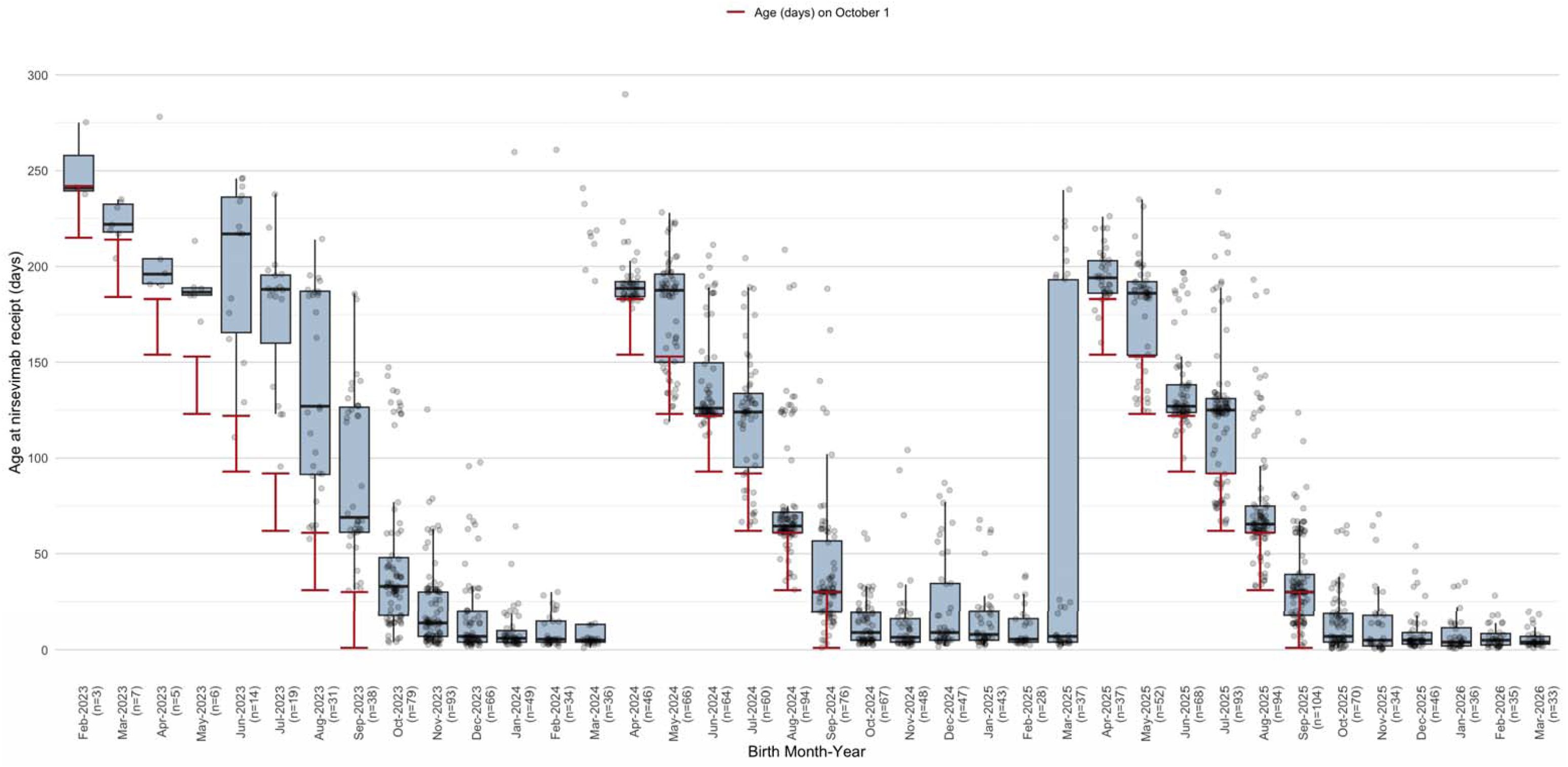
Age at nirsevimab receipt by birth month among infants who received nirsevimab Boxes show median and interquartile range of age at nirsevimab receipt by birth month. Points are individual infants. Red brackets indicate the range of ages on October 1 when infants born outside of the RSV season would be first eligible for nirsevimab. Two infants born in August 2023 who received their first documented nirsevimab dose during their second season at 477 and 578 days are not shown but were included in the boxplot statistics.

### Effectiveness against hospitalization

Among infants who received nirsevimab, five (0.3%) experienced an RSV hospitalization compared to 25 (0.8%) infants who did not receive nirsevimab (Table 1). At 180 days after receipt, nirsevimab reduced the risk of RSV hospitalization by 95% (95% CI: 84% to 98%) compared to no intervention across three seasons (Table 2, Figure 2A&B). Nirsevimab reduced the absolute risk of RSV hospitalization by 6.8 cases (95% CI: 3.0 to 10.6) per 1000 infants, for an NNI of 147 (95% CI: 94 to 333). Effectiveness against RSV hospitalization at 180-days post receipt was similar when adding a seven-day immune lag (RRR: 95% [95% CI: 84% to 98%]; RD: -6.6 per 1000 [95% CI: -10.4 to -2.8]) (Table S2).

**Figure 2.**
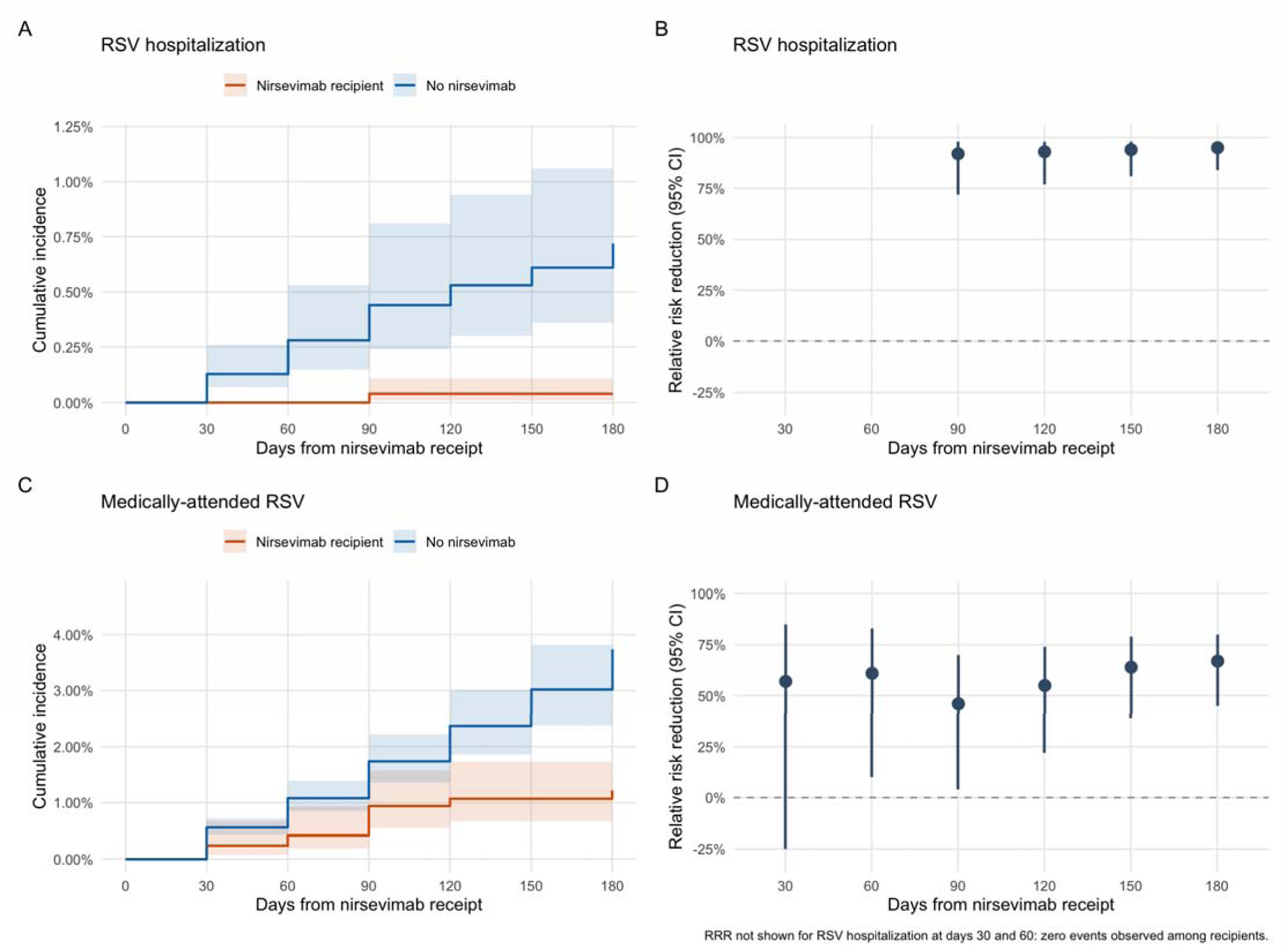
Cumulative incidence of RSV hospitalization and medically-attended RSV by nirsevimab receipt, and relative risk reduction, with 95% confidence intervals Panel A: Cumulative incidence of RSV hospitalization; B. Relative risk reduction against RSV hospitalization; C. Cumulative incidence of medically-attended RSV; D. Relative risk reduction against medically-attended RSV. Shaded bands and vertical bars are 95% confidence intervals from 1000 bootstrap samples. Estimates were computed at 30-day intervals from nirsevimab receipt and are drawn as steps between those time points. Abbreviations: RSV = respiratory syncytial virus; RRR = relative risk reduction

**Table 2.** Cumulative incidence of RSV hospitalization and medically-attended RSV by nirsevimab receipt and effectiveness estimates with 95% confidence intervals.

| Days from nirsevimab receipt | Cumulative incidence without nirsevimab per 1000 | Cumulative incidence with nirsevimab per 1000 | Relative risk reduction<br>(1 – RR) | Risk difference per 1000 | Number needed to immunize |
|---|---|---|---|---|---|
| **RSV hospitalization** |  |  |  |  |  |
| 30 | 1.3 (0.7 to 2.6) | 0.0 | NE | -1.3 (-2.2 to 0.4) | 769 (454 to 2500) |
| 60 | 2.8 (1.5 to 5.3) | 0.0 | NE | -2.8 (-4.6 to -1.0) | 357 (217 to 1000) |
| 90 | 4.4 (2.4 to 8.1) | 0.4 (0.1 to 1.0) | 92% (72% to 98%) | -4.1 (-6.9 to -1.2) | 243 (144 to 833) |
| 120 | 5.3 (3.0 to 9.4) | 0.4 (0.1 to 1.0) | 93% ( 77% to 98%) | -5.0 (-8.2 to -1.8) | 200 (121 to 555) |
| 150 | 6.1 (3.6 to 10.6) | 0.4 (0.1 to 1.0) | 94% (81% to 98%) | -5.8 (-9.2 to -2.4) | 172 (108 to 416) |
| 180 | 7.2 (4.3 to 12.0) | 0.4 (0.1 to 1.0) | 95% (84% to 98%) | -6.8 (-10.6 to -3.0) | 147 (94 to 333) |
| **Medically-attended RSV** |  |  |  |  |  |
| 30 | 5.6 (4.3 to 7.2) | 2.4 (0.8 to 6.7) | 57% (-25% to 85%) | -3.2 (-5.9 to -0.4) | 312 (169 to 2500) |
| 60 | 10.8 (8.5 to 13.9) | 4.2 (1.9 to 9.4) | 61% (10% to 83%) | -6.6 (-10.8 to -2.5) | 151 (92 to 400) |
| 90 | 17.4 (13.6 to 22.2) | 9.4 (5.5 to 15.8) | 46% (4% to 70%) | -8.0 (-14.4 to -1.6) | 125 (69 to 625) |
| 120 | 23.7 (18.6 to 30.1) | 10.7 (6.6 to 17.3) | 55% (22% to 74%) | -12.9 (-20.6 to -5.2) | 77 (48 to 192) |
| 150 | 30.2 (23.8 to 38.2) | 10.7 (6.6 to 17.3) | 64% (39% to 79%) | -19.4 (-28.4 to -10.5) | 51 (35 to 95) |
| 180 | 37.4 (29.5 to 47.3) | 12.2 (7.8 to 19.2) | 67% (45% to 80%) | -25.2 (-35.8 to -14.5) | 39 (28 to 68) |

NE: not estimable; no RSV hospitalizations occurred among nirsevimab recipients within 60 days of receipt. 95% confidence intervals are from 1000 nonparametric bootstrap samples. Relative risk reduction is shown as (1 – RR)\*100%.
Abbreviations: CI = confidence interval; NE = not estimable; RR = risk ratio; RSV = respiratory syncytial virus

The risk difference of bronchiolitis hospitalization among infants who received nirsevimab compared to infants who did not was -3.8 (95% CI: −10.1 to 2.4) per 1000 infants after 180 days of follow-up from receipt, for an NNI of 263 (95% CI: 99 to ∞; 416 to ∞) (Table S3). At 180 days from nirsevimab receipt, the risk difference of ARI hospitalization among infants who received nirsevimab compared to infants who did not was -4.9 (95% CI: −11.3 to 1.5) per 1000 infants, for an NNI of 204 (95% CI: 88 to ∞; 666 to ∞).

### Effectiveness against medically-attended illness

Among infants who received nirsevimab, 49 (2.6%) experienced medically-attended RSV (Table 1), including 27 outpatient visits, 13 emergency department visits, five hospitalizations, and four virtual care visits. Among infants who did not receive an intervention, 169 (5.7%) infants experienced medically-attended RSV, including 95 outpatient visits, 44 emergency department visits, 22 hospitalizations, and eight virtual care visits.

At 180 days from receipt, nirsevimab reduced the risk of medically-attended RSV by 67% (95% CI: 45% to 80%) compared to no intervention (Table 2, Figure 2C&D). Nirsevimab reduced the absolute risk of medically-attended RSV by 25.2 cases (95% CI: 14.5 to 35.8) per 1000 infants, for an NNI of 39 (95% CI: 28 to 68). Effectiveness against medically-attended RSV at 180-days post receipt was similar when adding a seven-day immune lag (RRR: 68% [95% CI: 44% to 92%]; RD: -24.8 per 1000 [95% CI: -35.0 to -14.5]) (Table S2).

Among infants born before 2025-2026 who could be followed for a second year, first dose nirsevimab effectiveness against the first medically-attended RSV episode was 65% (95% CI: 36% to 80%) at six months, 60% (95% CI: 35% to 75%) at one year, and 50% (95% CI: 24% to 67%) at two years after receipt (Table S4). The cumulative absolute reduction in first medically-attended RSV persisted through the second year; that is, the first-season benefit was not offset by later infections. Nirsevimab reduced the absolute risk of medically-attended RSV between 34.1 cases (95% CI: 14.4 to 53.9) and 47.3 cases (95% CI: 23.7 to 70.9) per 1000 infants during the second year post-receipt.

The risk difference of medically-attended bronchiolitis among infants who received nirsevimab compared to infants who did not was -15.3 (95% CI: −33.1 to 2.4) per 1000 infants after 180 days of follow-up from receipt, for an NNI of 65 (95% CI: 3 to ∞; 416 to ∞) (Table S3). At 180 days from nirsevimab receipt, the risk difference of medically-attended ARI among infants who received nirsevimab compared to infants who did not was 22.0 (95% CI: −12.5 to 56.5) per 1000 infants.

## Discussion

Among 4,826 mother-infant pairs followed across three RSV seasons, nirsevimab substantially reduced the risk of RSV hospitalization and medically-attended RSV. We estimated these effects using a cohort design with a target trial framework in contrast to the test-negative design that has predominantly been used to evaluate nirsevimab effectiveness to date.

A major strength of our analysis was the study design, which allowed us to assess nirsevimab effectiveness using an estimand aligned in follow-up time with the pivotal nirsevimab RCTs. Specifically, this estimand measured effectiveness from time of nirsevimab receipt rather than from birth or season start. Achieving this alignment between observational and RCT estimands commonly relies on matching-based approaches, which can compromise generalizability and efficiency. To avoid these tradeoffs, we applied a novel method for assigning a randomization time to infants who did not receive nirsevimab, allowing us to achieve this alignment without matching while still adjusting for measured confounders.

At 150 days, the RCTs for nirsevimab estimated a 77% (95% CI: 50% to 90%) pooled efficacy against hospitalization for RSV LRTI and 80% (95% CI: 66% to 88%) efficacy against medically-attended RSV-associated LRTI.^16^ In comparison, we observed that the effectiveness of nirsevimab against RSV hospitalization was 94% (95% CI: 81 to 98%) and medically-attended RSV among infants was 64% (95% CI: 39% to 79%) at 150 days post-intervention. Of note, our study did not specify that RSV-related outcomes were required to be LRTIs. Through the TTE framework, we improved the interpretability of results compared to studies using TND. Our hospitalization estimate also sits at the upper end of pooled post-licensure estimates, which cluster around 80–85% across more than 40 observational studies,^7,28,29^ and of the 2024–2025 U.S. test-negative estimates of 77–81%.^19,24^ Our estimate against medically-attended RSV (67%) matches a three-season test-negative analysis in Connecticut spanning 2023–2024 through 2025–2026 (67%; 89% against hospitalization), which also found lower effectiveness in later seasons as RSV activity shifted later in the year.^50^ The lower effectiveness against medically-attended RSV was also somewhat expected since that outcome includes outpatient and virtual visits coded without testing, so both misclassification and differential care-seeking may pull the estimate toward the null.

Another strength of our study relates to the underlying cohort. By leveraging a well-defined cohort within KPGA we mitigated the risk of collider bias from conditioning on testing^33^ since infants were not required to undergo testing to be included. This represents an improvement over TNDs, the predominant approach used to estimate nirsevimab effectiveness, which by design restricts the study population to infants who both sought healthcare and were tested. Because inclusion in our cohort was not restricted to infants who sought medical care, our design also improves generalizability and reduces the potential for selection bias. In addition, all mother-infant pairs were KPGA members with coverage and access to care through the same integrated system, limiting major differences in healthcare access and seeking behavior between nirsevimab recipients and non-recipients. Moreover, linking infants to their mothers’ records within KPGA allowed us to investigate important maternal covariates that impact infant health and the decision to immunize and achieve clearer documentation of maternal vaccination status.

This study design also allowed us to estimate additive effects, which offer a more direct measure of averted burden than relative effect estimates alone. Although the relative effectiveness of nirsevimab against medically-attended RSV was lower than that observed for RSV hospitalization, the corresponding additive effect was larger, reflecting the higher baseline incidence of medically-attended RSV (Table 2). Additive effects also enabled estimation of NNI, providing interpretable measures of prevention product impact. The NNI of 147 for RSV hospitalization is higher than estimates from universal programs in Galicia (NNI 25)^51^ and Ireland (NNI 54),^52^ which reflects the lower baseline hospitalization risk in our commercially insured cohort (0.8% among non-recipients over follow-up) and the claims-based outcome rather than lower effectiveness.

This study was subject to limitations. First, we relied on ICD-10 diagnostic codes rather than laboratory-confirmed diagnostic testing to identify RSV-specific outcomes. Because KPGA does not directly provide emergency department or inpatient care, diagnostic testing results from those settings were not available. Even among outpatient encounters, only 43% of those coded with an RSV ICD-10 diagnosis had a documented positive test result while the remainder had no test recorded. Importantly, no encounters coded with an RSV ICD-10 diagnosis were accompanied by a documented negative test result. This pattern raises the possibility of outcome misclassification. However, had we required laboratory confirmation as TNDs do, we may have excluded a substantial proportion of clinically diagnosed RSV cases given infrequent testing in this setting. To capture potentially undetected RSV, we additionally evaluated broader outcomes including all-cause bronchiolitis hospitalization, of which RSV is estimated to account for 60-80% of cases among infants,^53^ medically-attended bronchiolitis, all-cause ARI hospitalization, and medically-attended ARI. Point estimates for bronchiolitis and ARI hospitalization favored nirsevimab, but confidence intervals included the null (Table S3).

Next, our analysis may be affected by a data lag. We pulled data on June 16, 2026, shortly after the end of the study period on May 30, 2026. Records for the most recent months may have been incomplete at the time of extraction. Data lag likely also contributed to the lower proportion of infants linked to maternal records and to lower recorded nirsevimab receipt among the most recent births. To assess the influence of the data lag, we conducted an analysis ending follow-up on March 16, 2026, allowing a 90-day run-out period before data extraction. We found that effectiveness estimates were similar to the primary analysis (Table S5).

Second, although we adjusted for healthcare-seeking behavior using the number of prenatal encounters during pregnancy, maternal Tdap and influenza vaccine receipt during pregnancy, and race and all mother-infant pairs were part of the same integrated health system, residual confounding by care-seeking is possible: in some intervals (90 to 120 days after receipt) nirsevimab recipients had a higher absolute risk of medically-attended ARI (Table S3), which is more plausibly a marker of care-seeking than an effect of the antibody. Infants who received nirsevimab may have had mothers with a greater propensity to seek healthcare, as suggested by the higher rate of maternal vaccination for Tdap, influenza, and COVID-19 among mothers of nirsevimab recipients compared to mothers of infants who did not receive nirsevimab (Table 1). In contrast, we did not observe this pattern for all-cause ARI hospitalizations. Effectiveness estimates against hospitalization were less susceptible to this type of bias because severe cases may be likely to seek care regardless of nirsevimab status.

Additionally, the study population consisted exclusively of infants insured within KPGA. Although this improves comparability between nirsevimab recipients and non-recipients, it may limit the generalizability of our findings to uninsured infants or those receiving care in other settings. Insurance type is a likely determinant of uptake of RSV preventive interventions,^44^ which was not considered in this study. In addition, the sample size was insufficient to evaluate effectiveness by season, age, and births during or out of season, and whether effectiveness meaningfully changed over time within the season. We also could not evaluate clesrovimab, which became available in 2025– 2026, and our comparison group excluded infants protected by maternal vaccination; as maternal RSVpreF uptake grows, the unprotected pool shrinks and the population to which these estimates apply changes. Finally, we adjusted for baseline characteristics at birth or close to birth rather than include time-varying confounders.

Using a target trial framework and a novel method for assigning a randomization time to infants who did not receive nirsevimab, we measured nirsevimab effectiveness estimates that were less susceptible to the collider bias and restricted generalizability inherent to the test-negative designs and matching-based studies that have predominated. Notably, these real-world estimates were broadly consistent with the efficacy observed in phase 3 RCTs, demonstrating that target trial emulation with a clearly specified estimand can recover trial findings from observational data. With follow-up extending through the 2025–2026 RSV season, these findings provide timely, real-world evidence to support the continued use of nirsevimab in protecting infants against RSV disease. With two long-acting antibodies now recommended without preference,^11^ and with sporadic nirsevimab-resistant RSV-B variants reported in immunized children in Spain during 2025–2026,^54,55^ monitoring of effectiveness in integrated systems that include near real-time data such as ours remains important.

## Data Availability

The data underlying this study contain protected health information and cannot be shared publicly. De-identified data may be made available from the corresponding author upon reasonable request, subject to approval by Kaiser Permanente Georgia and execution of a data use agreement.

## Acknowledgements

Funding: This publication was made possible by the Insight Net cooperative agreement CDC-RFA-FT-23-0069 from the CDC’s Center for Forecasting and Outbreak Analytics. Its contents are solely the responsibility of the authors and do not necessarily represent the official views of the Centers for Disease Control and Prevention.

