## Supplemental Materials for "Real-world effectiveness of nirsevimab over three RSV seasons in an integrated health care system: a target trial emulation study"

Supplementary materials

Table S1. Target trial specification for nirsevimab compared to no intervention for the prevention of RSV hospitalization and medically-attended RSV among infants

| Protocol component | Target trial specification | Implementation with observational data |
| --- | --- | --- |
| Eligibility criteria | Inclusion: Infants born on or after February 2, 2023 and on or before May 30, 2026  Exclusion: Mother’s medical history during pregnancy is unknown or infant received palivizumab, or mother received RSVpreF vaccination ≥14 days before delivery | Inclusion: Same as target trial; born on or before May 30, 2026 (infants aged <8 months on October 1, 2023, through May 30, 2026)  Exclusion: Infants who   - Could not be paired to mother’s record - Not enrolled in KPGA within 32 days of birth, or not enrolled 33–75 days from birth without a KPGA healthcare encounter within 75 days of birth - Received palivizumab - Received an unknown RSV preventive product - Mother received RSVpreF vaccination ≥14 days before delivery |
| Intervention strategies | Nirsevimab or placebo | - First dose of nirsevimab at any point after birth - No receipt of nirsevimab |
| Intervention assignment | Randomized | Observed, but emulate randomization via standardization on infant age, maternal age at delivery, number of prenatal encounters during the relevant pregnancy, calendar time, preterm birth status, presence of congenital malformations of the circulatory system, presence of congenital pulmonary abnormalities, and presence of neuromuscular conditions. |
| Time zero | Nirsevimab or placebo receipt | Nirsevimab receipt for recipients and the assigned randomization time for non-recipients |
| Follow-up period | From nirsevimab receipt until the first of an outcome, loss to follow up, or end of follow-up at six months from nirsevimab receipt | From nirsevimab receipt until the first of an outcome, loss to follow up indicated by a lapse in KPGA coverage of more than 75 days, end of follow-up at six months from nirsevimab receipt, or last date of data collection (May 30, 2026). Follow-up among controls additionally ends at nirsevimab receipt. |
| Outcome | - Laboratory confirmed RSV hospitalization - Laboratory confirmed medically-attended RSV | - Hospitalization with RSV ICD-10 code - Medically-attended RSV illness, including virtual care visits, outpatient visits, emergency department visits, and hospitalizations, identified via ICD-10 code |
| Causal contrast | - Cumulative incidence of outcome at 30, 60, 90, 120, 150, and 180 days following nirsevimab receipt among nirsevimab recipients versus among infants who received placebo (ratio and difference) | - Cumulative incidence of outcome at 30, 60, 90, 120, 150, and 180 days following nirsevimab receipt among nirsevimab recipients versus among infants who have not received nirsevimab (ratio and difference) |
| Analysis plan | Kaplan-Meier survival function | G-computation with separate Cox models in recipients and non-recipients with age in days as the time scale, the adjustment set, and 1000 bootstrap samples for CIs |

Abbreviations: RSV = respiratory syncytial virus; KPGA = Kaiser Permanente Georgia; ICD = International Classification of Diseases; CI = confidence interval

Table S2. Cumulative incidence of RSV hospitalization and medically-attended RSV by nirsevimab receipt and effectiveness estimates including a seven-day immune lag with 95% confidence intervals

| **Days from nirsevimab receipt** | **Cumulative incidence without nirsevimab per 1000** | **Cumulative incidence with nirsevimab per 1000** | **Relative risk reduction** | **Risk difference per 1000** |
| --- | --- | --- | --- | --- |
| **RSV hospitalization** |  |  |  |  |
| 30 | 1.1 (0.6 to 2.1) | 0.0 | NE | -1.1 (-1.8 to -0.4) |
| 60 | 2.6 (1.3 to 5.1) | 0.0 | NE | -2.6 (-4.4 to -0.8) |
| 90 | 4.2 (2.2 to 7.9) | 0.4 (0.1 to 1.0) | 91% (71% to 97%) | -3.8 (-6.7 to -1.0) |
| 120 | 5.1 (2.8 to 9.3) | 0.4 (0.1 to 1.0) | 93% (77% to 98%) | -4.8 (-7.9 to -1.6) |
| 150 | 5.9 (3.4 to 10.4) | 0.4 (0.1 to 1.0) | 94% (80% to 98%) | -5.6 (-9.0 to -2.1) |
| 180 | 7.0 (4.1 to 12.0) | 0.4 (0.1 to 1.0) | 95% (84% to 98%) | -6.6 (-10.4 to -2.8) |
| **Medically-attended RSV** |  |  |  |  |
| 30 | 4.5 (3.5 to 5.9) | 1.8 (0.6 to 5.5) | 60% (-26% to 87%) | -2.7 (-5.2 to -0.3) |
| 60 | 9.6 (7.6 to 12.6) | 3.6 (1.4 to 9.1) | 63% (4% to 86%) | -6.2 (-10.0 to -2.2) |
| 90 | 16.4 (12.8 to 21.0) | 8.8 (5.0 to 15.5) | 46% (0% to 71%) | -7.6 (-13.8 to -1.4) |
| 120 | 22.7 (17.8 to 28.9) | 10.2 (6.0 to 17.2) | 55% (20% to 75%) | -12.5 (-20.0 to -5.0) |
| 150 | 29.2 (23.0 to 37.0) | 10.2 (6.0 to 17.2) | 65% (38% to 81%) | -19.0 (-27.7 to -10.4) |
| 180 | 36.4 (28.7 to 46.0) | 11.7 (7.0 to 19.2) | 68% (44% to 82%) | -24.8 (-35.0 to -14.5) |

No RSV hospitalizations occurred among nirsevimab recipients within 60 days of receipt. 95% confidence intervals are from 1000 nonparametric bootstrap samples. Relative risk reduction is shown as a proportion (1 − RR)*100%.

Abbreviations: CI = confidence interval; NE = not estimable; RR = risk ratio; RSV = respiratory syncytial virus

Table S3. Cumulative incidence of all-cause bronchiolitis and acute respiratory illness by nirsevimab receipt and effectiveness estimates with 95% confidence intervals

| **Days from nirsevimab receipt** | **Risk difference per 1000**  **(95% CIs)** | **Number needed to immunize (95% CIs)^a^** |
| --- | --- | --- |
| **Bronchiolitis hospitalization** |  |  |
| 30 | −1.2 (−3.2 to 0.9) | 833 (312 to $\infty$; 1111 to $\infty)$ |
| 60 | −1.3 (−4.8 to 2.2) | 769 (208 to $\infty$; 454 to $\infty)$ |
| 90 | −2.8 (−7.4 to 1.7) | 357 (135 to $\infty$; 588 to $\infty)$ |
| 120 | −4.5 (−9.4 to 0.4) | 222 (106 to $\infty$; 2500 to $\infty)$ |
| 150 | −3.8 (−9.5 to 1.9) | 263 (105 to $\infty$; 526 to $\infty)$ |
| 180 | −3.8 (−10.1 to 2.4) | 263 (99 to $\infty$; 416 to $\infty)$ |
| **Medically-attended bronchiolitis** |  |  |
| 30 | 0.5 (−5.0 to 6.1) | 2000 (200 to $\infty$; 163 to $\infty)$ |
| 60 | 0.9 (−7.4 to 9.2) | 1111 (135 to $\infty$; 108 to $\infty)$ |
| 90 | 0.0 (−11.3 to 11.2) | 32,009 (88 to $\infty$; 89 to $\infty)$ |
| 120 | −8.3 (−21.5 to 5.1) | 120 (46 to $\infty$; 196 to $\infty)$ |
| 150 | −10.3 (−26.1 to 5.4) | 97 (38 to $\infty$; 185 to $\infty)$ |
| 180 | −15.3 (−33.1 to 2.4) | 65 (30 to $\infty$; 416 to $\infty)$ |
| **ARI hospitalization** |  |  |
| 30 | −1.6 (−3.6 to 0.4) | 625 (277 to $\infty$; 2500 to $\infty)$ |
| 60 | −2.1 (−5.6 to 1.3) | 476 (178 to $\infty$; 769 to $\infty)$ |
| 90 | −3.0 (−7.6 to 1.6) | 333 (131 to $\infty$; 625 to $\infty)$ |
| 120 | −4.7 (−9.7 to 0.4) | 212 (103 to $\infty$; 2500 to $\infty)$ |
| 150 | −4.1 (−9.9 to 1.7) | 243 (101 to $\infty$; 588 to $\infty)$ |
| 180 | −4.9 (−11.3 to 1.5) | 204 (88 to $\infty$; 666 to $\infty)$ |
| **Medically-attended ARI** |  |  |
| 30 | 3.6 (−9.9 to 17.2) | 277 (101 to $\infty$; 58 to $\infty)$ |
| 60 | 17.9 (−1.3 to 37.2) | 55 (769 to $\infty$; 26 to $\infty)$ |
| 90 | 40.8 (15.6 to 66.0) | 24 (15 to 64) |
| 120 | 38.1 (9.5 to 66.8) | 26 (15 to 64) |
| 150 | 30.1 (−2.1 to 62.4) | 33 (476 to $\infty$; 16 to $\infty)$ |
| 180 | 22.0 (−12.5 to 56.5) | 45 (80 to $\infty$; 17 to $\infty)$ |

^a^Reported discontinuous intervals for number needed to immunize as discussed in Altman DG. Confidence intervals for the number needed to treat. *BMJ*. 1998;317(7168):1309-1312. Intervals are shown as NNI for benefit to ∞; NNI for harm to ∞. Where the risk difference is positive (higher risk among nirsevimab recipients), the value is a number needed to harm.

Abbreviations: CI = confidence interval; NNI = number needed to immunize; RSV = respiratory syncytial virus; ARI = acute respiratory illness

Table S4. Relative risk reduction and risk difference of medically-attended RSV through 720 days post-intervention among infants born prior to the 2025-2026 season

| **Days from nirsevimab receipt** | **Relative risk reduction (95% CIs)** | **Risk difference per 1000 infants (95% CIs)** |
| --- | --- | --- |
| 30 | 42% (−65% to 80%) | −2.5 (−6.1 to 1.1) |
| 60 | 51% (−24% to 81%) | −5.4 (−10.4 to −0.5) |
| 90 | 38% (−19% to 67%) | −6.4 (−13.6 to 0.7) |
| 120 | 49% (6% to 73%) | −11.3 (−19.5 to −3.1) |
| 150 | 60% (26% to 78%) | −17.3 (−26.4 to −8.2) |
| 180 | 65% (36% to 80%) | −22.8 (−33.1 to −12.6) |
| 210 | 67% (42% to 82%) | −28.0 (−39.5 to −16.5) |
| 240 | 68% (44% to 81%) | −30.7 (−43.0 to −18.3) |
| 270 | 66% (42% to 80%) | −32.5 (−46.2 to −18.9) |
| 300 | 62% (37% to 77%) | −32.9 (−47.8 to −18.0) |
| 330 | 62% (37% to 77%) | −34.6 (−50.4 to −18.9) |
| 360 | 60% (35% to 75%) | −35.7 (−52.2 to −19.1) |
| 390 | 54% (29% to 71%) | −34.5 (−52.3 to −16.7) |
| 420 | 53% (27% to 69%) | −35.1 (−53.8 to −16.3) |
| 450 | 49% (22% to 66%) | −34.1 (−53.9 to −14.4) |
| 480 | 49% (23% to 66%) | −35.7 (−56.2 to −15.1) |
| 510 | 49% (23% to 66%) | −36.5 (−57.7 to −15.4) |
| 540 | 48% (23% to 66%) | −37.7 (−59.6 to −15.9) |
| 570 | 51% (26% to 67%) | −41.1 (−63.6 to −18.6) |
| 600 | 52% (27% to 68%) | −43.0 (−65.7 to −20.2) |
| 630 | 53% (29% to 68%) | −44.5 (−67.5 to −21.5) |
| 660 | 53% (30% to 69%) | −46.2 (−69.5 to −22.8) |
| 690 | 54% (31% to 69%) | −47.3 (−70.9 to −23.7) |
| 720 | 50% (24% to 67%) | −44.7 (−69.9 to −19.4) |

Estimates are cumulative from the time of nirsevimab receipt. They are not season-specific estimates of effectiveness during the second RSV season.

Abbreviations: RSV = respiratory syncytial virus; CI = confidence interval

Table S5. Cumulative incidence of RSV hospitalization and medically-attended RSV by nirsevimab receipt and effectiveness estimates with the analysis ending follow-up on March 16, 2026 for a 90 day run out period

| **Days from nirsevimab receipt** | **Cumulative incidence without nirsevimab** | **Cumulative incidence with nirsevimab** | **Relative risk reduction** | **Risk difference per 1000** |
| --- | --- | --- | --- | --- |
| **RSV hospitalization** |  |  |  |  |
| 30 | 1.5 (0.8 to 2.7) | 0.0 | NE | -1.5 (-2.4 to -0.5) |
| 60 | 3.2 (1.7 to 5.8) | 0.0 | NE | -3.2 (-5.2 to -1.1) |
| 90 | 5.1 (2.7 to 9.2) | 0.4 (0.1 to 1.1) | 93% (75% to 98%) | -4.7 (-7.9 to -1.4) |
| 120 | 6.1 (3.4 to 10.9) | 0.4 (0.1 to 1.1) | 94% (79% to 98%) | -5.7 (-9.4 to -2.1) |
| 150 | 7.1 (4.0 to 12.4) | 0.4 (0.1 to 1.1) | 95% (82% to 99%) | -6.7 (-10.8 to -2.7) |
| 180 | 8.4 (4.8 to 14.5) | 0.4 (0.1 to 1.1) | 96% (85% to 99%) | -8.0 (-12.7 to -3.4) |
| **Medically-attended RSV** |  |  |  |  |
| 30 | 6.2 (4.9 to 8.1) | 2.5 (0.9 to 7.1) | 60% (-19% to 86%) | -3.8 (-6.8 to -0.8) |
| 60 | 12.3 (9.6 to 15.7) | 4.5 (1.8 to 10.9) | 63% (13% to 84%) | -7.8 (-12.4 to -3.2) |
| 90 | 19.9 (15.6 to 25.3) | 10.2 (6.0 to 17.3) | 48% (10% to 70%) | -9.6 (-16.6 to -2.7) |
| 120 | 27.2 (21.3 to 34.5) | 11.8 (7.3 to 19.2) | 56% (24% to 74%) | -15.3 (-23.8 to -6.8) |
| 150 | 34.8 (27.3 to 44.2) | 12.9 (8.0 to 20.9) | 62% (35% to 78%) | -21.8 (-32.0 to -11.7) |
| 180 | 43.4 (33.9 to 55.5) | 15.7 (9.1 to 27.2) | 63% (33% to 80%) | -27.7 (-41.7 to -13.6) |

No RSV hospitalizations occurred among nirsevimab recipients within 60 days of receipt. 95% confidence intervals are from 1000 nonparametric bootstrap samples. Relative risk reduction is shown as a proportion (1 − RR).

Abbreviations: CI = confidence interval; NE = not estimable; RR = risk ratio; RSV = respiratory syncytial virus

Figure S1. Mother-infant paired cohort selection flowchart

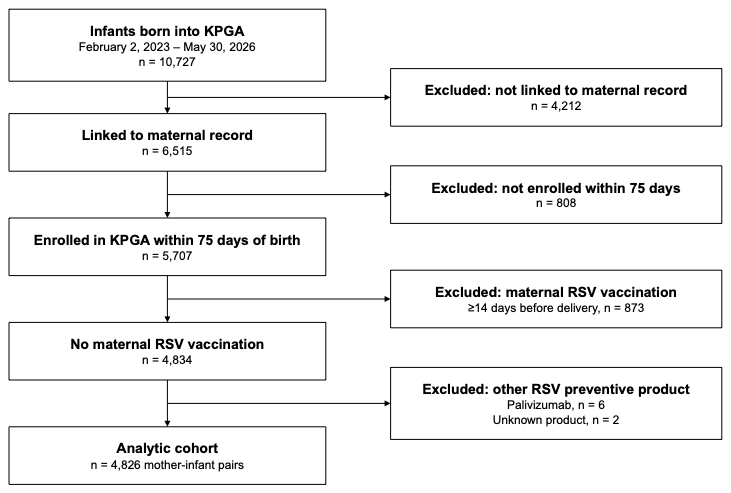

Abbreviations: KPGA = Kaiser Permanente Georgia; RSV = respiratory syncytial virus

Appendix A. ICD-10 codes for the identification of all-cause acute respiratory illness medical encounters.

| Code | Description |
| --- | --- |
| A37.00 | Whooping cough due to Bordetella pertussis, without pneumonia |
| A37.01 | Whooping cough due to Bordetella pertussis, with pneumonia |
| A37.10 | Whooping cough due to Bordetella parapertussis, without pneumonia |
| A37.11 | Whooping cough due to Bordetella parapertussis, with pneumonia |
| A37.80 | Whooping cough due to other Bordetella species, without pneumonia |
| A37.81 | Whooping cough due to other Bordetella species, with pneumonia |
| A37.90 | Whooping cough, unspecified species, without pneumonia |
| A37.91 | Whooping cough, unspecified species, with pneumonia |
| B25.0 | Cytomegaloviral pneumonitis |
| B97.4 | Respiratory syncytial virus as the cause of diseases classified elsewhere |
| J00 | Acute nasopharyngitis [common cold] |
| J01.00 | Acute maxillary sinusitis, unspecified |
| J01.01 | Acute recurrent maxillary sinusitis |
| J01.10 | Acute frontal sinusitis, unspecified |
| J01.11 | Acute recurrent frontal sinusitis |
| J01.20 | Acute ethmoidal sinusitis, unspecified |
| J01.21 | Acute recurrent ethmoidal sinusitis |
| J01.30 | Acute sphenoidal sinusitis, unspecified |
| J01.31 | Acute recurrent sphenoidal sinusitis |
| J01.40 | Acute pansinusitis, unspecified |
| J01.41 | Acute recurrent pansinusitis |
| J01.80 | Other acute sinusitis |
| J01.81 | Other acute recurrent sinusitis |
| J01.90 | Acute sinusitis, unspecified |
| J01.91 | Acute recurrent sinusitis, unspecified |
| J02.0 | Streptococcal pharyngitis |
| J02.8 | Acute pharyngitis due to other specified organisms |
| J02.9 | Acute pharyngitis, unspecified |
| J03.00 | Acute streptococcal tonsillitis, unspecified |
| J03.01 | Acute recurrent streptococcal tonsillitis |
| J03.80 | Acute tonsillitis due to other specified organisms |
| J03.81 | Acute recurrent tonsillitis due to other specified organisms |
| J03.90 | Acute tonsillitis, unspecified |
| J03.91 | Acute recurrent tonsillitis, unspecified |
| J04.0 | Acute laryngitis |
| J04.10 | Acute tracheitis without obstruction |
| J04.11 | Acute tracheitis with obstruction |
| J04.2 | Acute laryngotracheitis |
| J04.30 | Supraglottitis, unspecified, without obstruction |
| J04.31 | Supraglottitis, unspecified, with obstruction |
| J05.0 | Acute obstructive laryngitis [croup] |
| J05.10 | Acute epiglottitis without obstruction |
| J05.11 | Acute epiglottitis with obstruction |
| J06.0 | Acute laryngopharyngitis |
| J06.9 | Acute upper respiratory infection, unspecified |
| J09.X1 | Influenza due to identified novel influenza A virus with pneumonia |
| J09.X2 | Influenza due to identified novel influenza A virus with other respiratory manifestations |
| J09.X3 | Influenza due to identified novel influenza A virus with gastrointestinal manifestations |
| J09.X9 | Influenza due to identified novel influenza A virus with other manifestations |
| J10.00 | Influenza due to other identified influenza virus with unspecified type of pneumonia |
| J10.01 | Influenza due to other identified influenza virus with the same other identified influenza virus pneumonia |
| J10.08 | Influenza due to other identified influenza virus with other specified pneumonia |
| J10.1 | Influenza due to other identified influenza virus with other respiratory manifestations |
| J10.2 | Influenza due to other identified influenza virus with gastrointestinal manifestations |
| J10.81 | Influenza due to other identified influenza virus with encephalopathy |
| J10.82 | Influenza due to other identified influenza virus with myocarditis |
| J10.83 | Influenza due to other identified influenza virus with otitis media |
| J10.89 | Influenza due to other identified influenza virus with other manifestations |
| J11.00 | Influenza due to unidentified influenza virus with unspecified type of pneumonia |
| J11.08 | Influenza due to unidentified influenza virus with specified pneumonia |
| J11.1 | Influenza due to unidentified influenza virus with other respiratory manifestations |
| J11.2 | Influenza due to unidentified influenza virus with gastrointestinal manifestations |
| J11.81 | Influenza due to unidentified influenza virus with encephalopathy |
| J11.82 | Influenza due to unidentified influenza virus with myocarditis |
| J11.83 | Influenza due to unidentified influenza virus with otitis media |
| J11.89 | Influenza due to unidentified influenza virus with other manifestations |
| J12.0 | Adenoviral pneumonia |
| J12.1 | Respiratory syncytial virus pneumonia |
| J12.2 | Parainfluenza virus pneumonia |
| J12.3 | Human metapneumovirus pneumonia |
| J12.81 | Pneumonia due to SARS-associated coronavirus |
| J12.89 | Other viral pneumonia |
| J12.9 | Viral pneumonia, unspecified |
| J13 | Pneumonia due to Streptococcus pneumoniae |
| J14 | Pneumonia due to Hemophilus influenzae |
| J15.0 | Pneumonia due to Klebsiella pneumoniae |
| J15.1 | Pneumonia due to Pseudomonas |
| J15.20 | Pneumonia due to staphylococcus, unspecified |
| J15.211 | Pneumonia due to Methicillin susceptible Staphylococcus aureus |
| J15.212 | Pneumonia due to Methicillin resistant Staphylococcus aureus |
| J15.29 | Pneumonia due to other staphylococcus |
| J15.3 | Pneumonia due to streptococcus, group B |
| J15.4 | Pneumonia due to other streptococci |
| J15.5 | Pneumonia due to Escherichia coli |
| J15.6 | Pneumonia due to other Gram-negative bacteria (non-billable parent code) |
| J15.61 | Pneumonia due to Acinetobacter baumannii |
| J15.69 | Pneumonia due to other Gram-negative bacteria |
| J15.7 | Pneumonia due to Mycoplasma pneumoniae |
| J15.8 | Pneumonia due to other specified bacteria |
| J15.9 | Unspecified bacterial pneumonia |
| J16.0 | Chlamydial pneumonia |
| J16.8 | Pneumonia due to other specified infectious organisms |
| J17 | Pneumonia in diseases classified elsewhere |
| J18.0 | Bronchopneumonia, unspecified organism |
| J18.1 | Lobar pneumonia, unspecified organism |
| J18.2 | Hypostatic pneumonia, unspecified organism |
| J18.8 | Other pneumonia, unspecified organism |
| J18.9 | Pneumonia, unspecified organism |
| J20.0 | Acute bronchitis due to Mycoplasma pneumoniae |
| J20.1 | Acute bronchitis due to Hemophilus influenzae |
| J20.2 | Acute bronchitis due to streptococcus |
| J20.3 | Acute bronchitis due to coxsackievirus |
| J20.4 | Acute bronchitis due to parainfluenza virus |
| J20.5 | Acute bronchitis due to respiratory syncytial virus |
| J20.6 | Acute bronchitis due to rhinovirus |
| J20.7 | Acute bronchitis due to echovirus |
| J20.8 | Acute bronchitis due to other specified organisms |
| J20.9 | Acute bronchitis, unspecified |
| J21.0 | Acute bronchiolitis due to respiratory syncytial virus |
| J21.1 | Acute bronchiolitis due to human metapneumovirus |
| J21.8 | Acute bronchiolitis due to other specified organisms |
| J21.9 | Acute bronchiolitis, unspecified |
| J22 | Unspecified acute lower respiratory infection |
| J39.8 | Other specified diseases of upper respiratory tract |
| J39.9 | Disease of upper respiratory tract, unspecified |
| J40 | Bronchitis, not specified as acute or chronic |
| J45.20 | Mild intermittent asthma, uncomplicated |
| J45.21 | Mild intermittent asthma with (acute) exacerbation |
| J45.22 | Mild intermittent asthma with status asthmaticus |
| J45.30 | Mild persistent asthma, uncomplicated |
| J45.31 | Mild persistent asthma with (acute) exacerbation |
| J45.32 | Mild persistent asthma with status asthmaticus |
| J45.40 | Moderate persistent asthma, uncomplicated |
| J45.41 | Moderate persistent asthma with (acute) exacerbation |
| J45.42 | Moderate persistent asthma with status asthmaticus |
| J45.50 | Severe persistent asthma, uncomplicated |
| J45.51 | Severe persistent asthma with (acute) exacerbation |
| J45.52 | Severe persistent asthma with status asthmaticus |
| J45.901 | Unspecified asthma with (acute) exacerbation |
| J45.902 | Unspecified asthma with status asthmaticus |
| J45.909 | Unspecified asthma, uncomplicated |
| J45.990 | Exercise induced bronchospasm |
| J45.991 | Cough variant asthma |
| J45.998 | Other asthma |

Abbreviations: ICD = International Classification of Diseases, 10th Revision
